# Practice and challenges of newborn hearing screening: Analysis of a five-year database in Italy

**DOI:** 10.1101/2025.10.15.25338079

**Authors:** Davide Coraci, Marta Fantoni, Eleonora Tonon, Raffaella Marchi, Luca Ronfani, Eva Orzan

**Affiliations:** Institute for Maternal and Child Health IRCCS Burlo Garofolo Trieste, Italy

## Abstract

Early identification and treatment of hearing impairments are essential for children’s development. International guidelines recommend a stepwise approach for conductive hearing screening in newborns. However, while the majority of countries around the world implemented universal newborn hearing screening, inconsistencies remain in terms of procedures and data management.

In particular, Level 3 of the screening pathway—comprising diagnostic confirmation and therapeutic management—has received limited attention in the literature, despite its central role in determining program effectiveness and patient outcomes.

This study investigates the clinical and organizational aspects of Level 3 within the neonatal hearing screening program of the Friuli-Venezia Giulia Region in Italy, analyzing data from 106 children enrolled between 2019 and 2023. The analysis considers the regional protocol, the roles of birthing centers, pediatricians, hospitals, and the Regional Center for Pediatric Hearing Loss Care, and subdivides Level 3 into four Phases (A–D) reflecting both organizational and diagnostic functions. By examining patient flow, false positives, loss to follow-up/documentation, and management practices, the study highlights how organizational factors—particularly the coordination between local and specialized facilities— produce “cascade” outcomes directly affecting diagnostic timelines and treatment initiation.

Findings provide critical insights into weaknesses of the current system and propose directions for improving program efficiency, accuracy, and overall quality of care.

## Introduction

Early diagnosis and treatment of congenital hearing impairment are fundamental for the development of perceptual and cognitive abilities, language, and overall social well-being. Nowadays, in many countries around the world, early intervention is made possible through established newborn hearing screening programs (Hyde, 2005). These programs follow the guidelines of the Joint Committee on Infant Hearing (JCIH), which define the key milestones and optimal timeline for the screening and management of suspected hearing loss in children, i.e., Early Hearing Detection and Intervention principles. Specifically, the JCIH (2019) recommends that screening programs include the following steps: an initial objective hearing screening and re-screening to be completed within the first month of life for all newborns (Level 1); a comprehensive audiological evaluation by no later than 2 months of age for infants who did not pass the initial screening (Level 2); confirmation of diagnosis and initiation of early intervention and treatment no later than 3 months of age (Level 3); and, finally, the implementation of an individualized audiological follow-up pathway for at-risk children. These guidelines update the 1-3-6 benchmark recommended in JCIH (2007).

In the case of Italy—the case study of this article—newborn hearing screening is mandatory and provided free of charge to all newborns. Even prior to 2017—the year of the most recent updates to national healthcare programs (Ministerial Decree *“Definition and Updating of the Essential Levels of Care,”* Art. 38)—the Italian clinical and scientific community was attentive and prepared to adapt its practices into large-scale, multidisciplinary, and multi-professional healthcare initiatives such as newborn hearing screening. However, the newborn hearing screening programs currently implemented across Italian regions are not without criticism. For example, in 2021, a report by the Italian Society of Otorhinolaryngology and Cervico-Facial Surgery (Orzan et al. 2022) highlights that the procedures and workflows of screening programs are not always consistent at the national, inter-regional, and sometimes even intra-regional levels. Additionally, there is a lack of systematic data analysis that could serve as process indicators for monitoring and improving the quality, accuracy, and time-efficiency of these programs (Holster et al. 2009; Walker et al. 2014; Orzan et al. 2016).

In particular, despite the existing literature on hearing screening programs (Kumar, Mohapatra 2011; Lim et al. 2012; Kanji et al. 2018; Marinho et al. 2020), limited attention is generally given to the later stages of these programs—stages in which the impact on the overall quality and effectiveness of the screening process can be appreciated not only for clinical and diagnostic elements, but also for organizational factors (Wen and Huang, 2023). While several publications have assessed the performance of the initial phases of hearing screening in Italy, namely, Levels 1 and 2 (Molini et al., 2016; Malesci et al., 2022), the subsequent stage, i.e., Level 3, which involves diagnostic confirmation and therapeutic management by a specialized facility, remains underexplored in the scientific literature.

However, assessing this final level of the hearing screening program is mandatory. It is at this stage that the most critical challenges often arise, as it is the point where the diagnosis is confirmed and treatment planning begins. Furthermore, evaluating Level 3 provides valuable insights into patient management, such as estimating the rates of false positives, lost to follow-up (LF), and lost documentation (LD) (Feresin et al. 2019; Orzan et al. 2021; Razak et al. 2021), and highlights how both clinical and organizational factors, including the coordination between local centers and specialized facilities, influence the overall effectiveness of the screening program.

The aim of this article is to examine the main clinical and organizational aspects involved in Level 3 of the hearing screening pathway, with the goal of identifying key management weaknesses, evaluating their cascade effects on diagnostic timelines and treatment initiation, and proposing potential improvements for future screening strategies. In particular, the article focuses on the Friuli Venezia Giulia (FVG) 2019-2023 Database (see Methods for details) of clinical records of 106 children that accessed the Level 3 of the neonatal hearing screening program, i.e., did not pass Level 1 examinations at birthing centers and the audiological evaluation administered during Level 2, in the Italian Region Friuli Venezia Giulia between 2019 and 2023.

FVG is a region in which the program for early identification of childhood hearing loss was formally established in 2012 (Feresin et al. 2019). In particular, the regional protocol for early identification of childhood hearing loss in FVG defines the roles of the different centers involved in the program and the procedures to follow. These include the different birthing centers in the region, responsible for performing the hearing screening test within the first month of life (Level 1), family pediatricians, responsible for hearing surveillance up to the child’s third year of life, three main hospitals (here indicated as Center 1, Center 2, and Center 3), responsible for conducting a complete audiological evaluation (Level 2), and the Regional Center for Pediatric Hearing Loss Care, the only one in the FVG region, located at the Burlo Garofolo Institute (Center 2), responsible for the final audiological and etiological diagnosis (Level 3), treatment, and follow-up of children with permanent hearing loss.

Data available in the dataset allows us to further analyze patients based on the different subsequent Phases 3A, 3B, 3C, and 3D (Figure 1) they encounter during Level 3. This subdivision permits to discriminate between the impact of clinical and organizational factors on the procedure as a whole, given the specific function each phase has. For instance, Phases 3A and 3B have primary organizational relevance, but a limited role from a diagnostic point of view, as they respectively concern individuals who failed Level 2 with suspected sensorineural or mixed hearing loss and individuals that correctly referred to Level 3 at the regional center for childhood hearing loss. Instead, Phases 3C and 3D, regarding respectively patients effectively arriving and evaluated at the regional center and patients with confirmed hearing losses that entered the diagnostic process, have clear diagnostic importance. From the analysis of the pathway followed by patients suspected of hearing loss, we are then able to assess the coverage of the screening hearing program from identification to diagnosis in FVG and, especially, assess the often overlooked impact of management and organizational factors on the efficiency of the program, formulating potential solutions to address observed issues.

**Figure 1.**
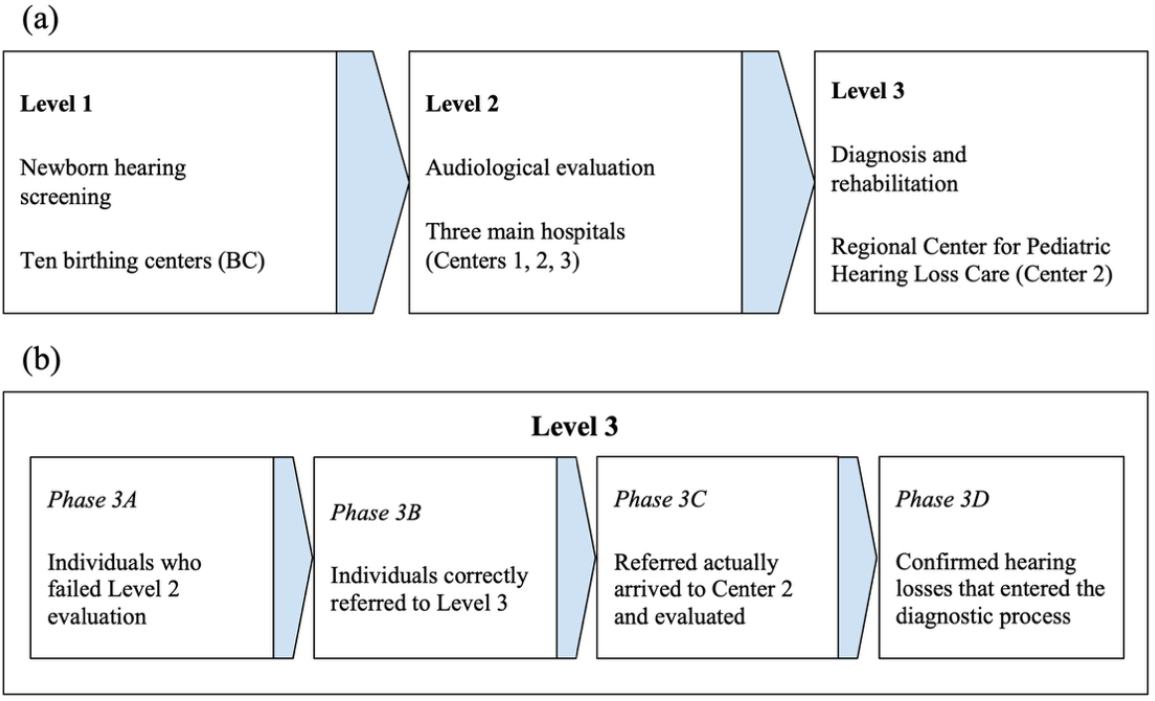
(a) Levels 1, 2, and 3 in the neonatal hearing screening program in the FVG region. (b) Details about the Level 3 Phases.

## Results

### 1.1 Overall screening procedure during Level 3 (Descriptive)

The records of 106 children who accessed Level 3 of the neonatal hearing screening program in FVG between 2019 and 2023 were analyzed through Phases 3A, 3B, 3C and 3D, until the final diagnosis, i.e., normal hearing condition (NORM), temporary (conductive) hearing loss subsequently solved (TT), and permanent (sensorineural, conductive or mixed) hearing loss (IP). Table 1 below reports the number of patients in the FVG region accessing Level 3, arranged by year, Phase, and final diagnosis (either temporary conducive hearing loss or permanent). Aggregated data are reported in a flowchart (Figure 2) below.

**Table 1.** Number of patients in the FVG region accessing Level 3 of the screening program between 2019 and 2023 and diagnosed with either TT or IP.

|  | Phase 3A | Phase 3B | Phase 3C | Phase 3D | TT | IP |
| --- | --- | --- | --- | --- | --- | --- |
| 2019 | 25 | 16 | 16 | 12 | 3 | 9 |
| 2020 | 19 | 16 | 15 | 13 | 1 | 12 |
| 2021 | 21 | 15 | 16 | 16 | 3 | 13 |
| 2022 | 23 | 20 | 21 | 19 | 3 | 16 |
| 2023 | 18 | 16 | 16 | 15 | 0 | 15 |
| Total | 106 | 83 | 84 | 75 | 10 | 65 |

**Figure 2.**
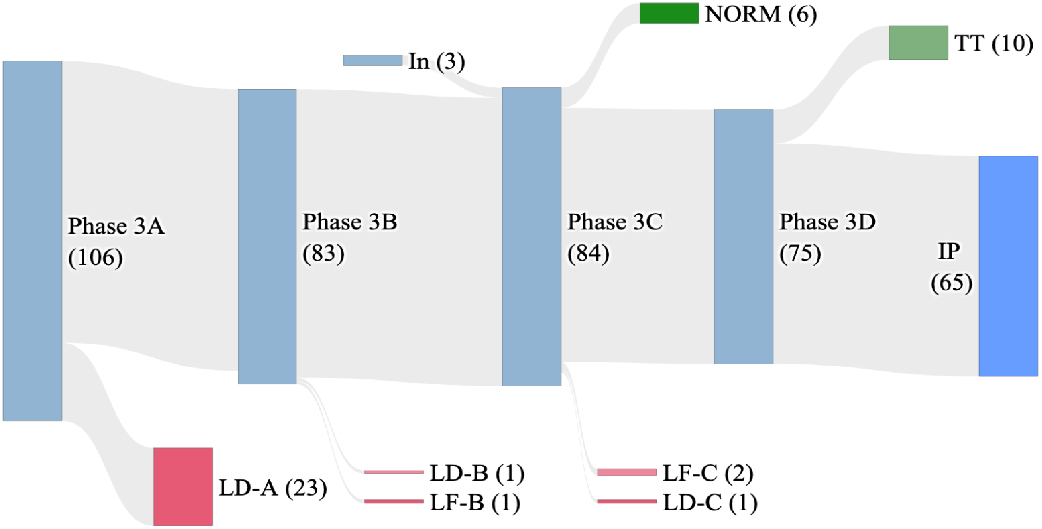
Flowchart of Level 3 showing aggregated data (2019–2023).

Among 106 patients accessing Level 3, only 65 were actually diagnosed as IP, while 10 patients resulted in TT and 6 NORM (See Supplementary materials). Across the different Phases, 28 records were classified as losses, specifically either lost documentation (LD), i.e., patients whose records are missing, or loss to follow-up (LF), i.e., patients enrolled in the program who are no longer in contact with the centers. Moreover, 3 patients (indicated as “In”) accessed Phase 3C directly, having them previously wrongly dismissed from the program.

Figure 2 shows mixed results. Indeed, while about 8 patients out of 10 correctly end the program with a definitive diagnosis and etiology, rates of LD and LF, as well as the presence of “In” patients reintroduced in the program, represent non-transcurable information when the efficiency of the screening procedure as a whole is evaluated.

Further insights about losses can be obtained by looking at the specific influence of organizational aspects on the screening program. In this respect, data grouped by the different hospitals, i.e., Center 1, 2, and 3, in charge of audiological assessment during Level 2, are relevant. Indeed, as shown in Figure 3, while IP, TT, and NORM are comparable between the three centers, it is worth noticing that all LD come from either Center 1 or 3, that is, centers other than the Regional Center for Pediatric Hearing Loss Care (Center 2) that treat patients during Level 2 and communicate referrals to the regional hub. Such data suggest a management issue when patients are referred from these centers to Center 2 that needs further investigation.

**Figure 3.**
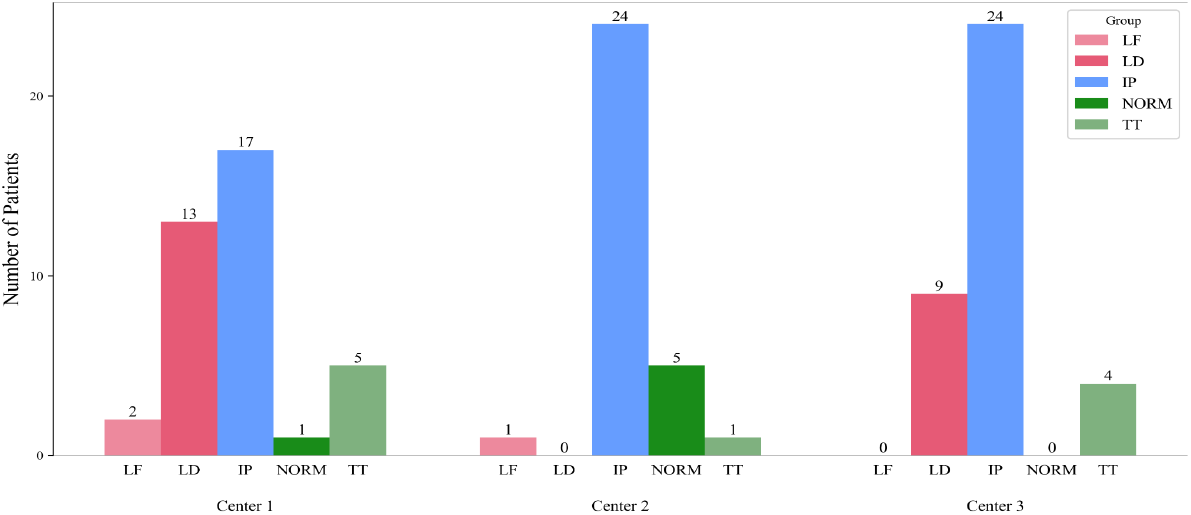
Outcomes (IP, TT, NORM, LD, and LF) are divided by Level 2 Centers (2019–2023).

### 1.2 Overall screening procedure during Level 3 (Statistical analysis)

Criticisms about the management of referrals, i.e., the high rate of losses, can be investigated in light of the specific functions that Phases of the program have. Indeed, Phases 3A to 3D in Level 3 have different organizational and clinical relevance. For instance, early Phases 3A and 3B have mainly organizational and management functions, as substantially related to the correct communication between the centers involved in the program, while Phases 3C and 3D have a clear clinical and diagnostic impact, as involving audiological evaluation, diagnosis, and treatment of patients.

Therefore, variations in the number of suspected hearing losses are not expected during Phases 3A and 3B and, if present, might be interpreted as cues of organizational and management weaknesses in the program. On the contrary, variations in the number of patients can be expected during Phases 3C and 3D, e.g., concerning the identification of false positives, given the clinical evaluations children undergo in these cases.

A straightforward way to evaluate the efficiency of the procedure and the influence of clinical and organizational Phases consists of estimating the incidence of hearing loss at each Phase. Given the total births in FVG during the five-year period 2019-2023 are 39,112, the overall incidence of hearing loss across the three Centers varies as follows: 2.71‰ (Phase 3A), 2.12‰ (Phase 3B), 2.15‰ (Phase 3C), 1.92‰ (Phase 3D). Then, the final incidences of temporary (TT) and permanent (IP) hearing impairments are respectively 0.26‰, and 1.66‰–results that, in the case of permanent hearing loss, are in line with the prevalence of the condition in Italy.

These results are in clear contrast with expectations about the screening program since the incidence varies during all Level 3 Phases. However, while variations are expected only during clinically-relevant Phases 3C and 3D, changes in the number of suspected hearing losses are observed during Phases 3A and 3B, that is, phases with primary organizational functions, e.g., concerning the correct communication of referrals between the centers involved in the program. In fact, these variations are due to the number of LF and LD only.

To further corroborate the hypothesis of some organizational issue at these stages of the program, data have been compared to the number of births (alive) in the FVG region over the 2019-2023 period, organized by birthing centers and Level 2 centers of reference (Table 2). This analysis allows us to have a detailed view of the local situations and, especially, to rule out causes other than organizational ones, such as the different number of births managed by each center.

**Table 2.** Number of births (alive) in the FVG region over the 2019-2023 period managed by birthing centers (BC) and aggregated by Level 2 centers of reference.

| Level 2 centers | Center 1 |  |  | Center 2 |  |  |  | Center 3 |  |  | Total year |
| --- | --- | --- | --- | --- | --- | --- | --- | --- | --- | --- | --- |
| Birthing centers | BC 1 | BC 2 | BC 3 | BC 4 | BC 5 | BC 6 | BC 7 | BC 8 | BC 9 | BC 10 |  |
| 2019 | 1,176 | 672 | 681 | 1,412 | 704 | 3 | 616 | 1,472 | 818 | 384 | 7,998 |
| 2020 | 1,190 | 498 | 556 | 1,475 | 826 | 0 | 537 | 1,537 | 836 | 390 | 7,918 |
| 2021 | 1,117 | 761 | 528 | 1,443 | 783 | 3 | 465 | 1,485 | 782 | 370 | 7,788 |
| 2022 | 1,147 | 699 | 510 | 1,563 | 829 | 11 | 503 | 1,505 | 724 | 311 | 7,866 |
| 2023 | 1,084 | 705 | 455 | 1,416 | 827 | 1 | 472 | 1,580 | 655 | 293 | 7,542 |
| Total births BC | 5,714 | 3,335 | 2,730 | 7,309 | 3,969 | 18 | 2,593 | 7,579 | 3,815 | 1,748 |  |
| Total births Center | 11,779 |  |  | 13,889 |  |  |  | 13,142 |  |  | 39,112 |

The percentage of suspected hearing impairments varies across centers. Considering the overall number of suspected hearing loss during Phase 3A, consisting in the total number of outcomes for each center reported in Fig. 3, i.e., 38 in Center 1, 31 in Center 2, and 37 in Center 3, and births effectively managed by each center, Center 1 counts an incidence of suspected hearing issues of 3.23‰ patients, while this percentage is equal to 2.23‰ and 2.82‰ for Centers 2 and 3, respectively. Instead, when the numbers of diagnosed IP after Phase 3D are considered (17 for Center 1 and 24 for both Center 2 and Center 3), these percentages indicate the actual incidence of IP, that is, 1.44‰ (Center 1), 1.73‰ (Center 2), and 1.83‰ (Center 3). These results are in line with the observed incidence of permanent hearing impairments in Italy; however, it is worth noting the difference between the incidence of Center 1 and the other two centers, in particular Center 3, which is the other Level 3 center that is not the regional hub.

As mentioned above, the lower incidence of IP from Center 1 as compared with other centers might be due to the lower number of total births administered by Center 1 between 2019 and 2023. In particular, given that births managed by Center 1 are much fewer than those administered by the other two centers, this might have biased the rate of patients accessing Level 3 coming from Center 1, the incidence of IP, and, in turn, our hypothesis that some organizational issues have impacted the management of referrals. However, this potential effect can be effectively ruled out by noting that the number of patients with suspected hearing impairments referred from Center 1 at Phase 3A is similar to that from Center 3 (38 vs. 37). Therefore, the difference in the number of births at Center 1 impacts only the incidence of suspected hearing loss at the very beginning of Level 3 (Phase 3A). Such an effect is, in fact, reduced over the subsequent phases of the program, where the incidence of IP in Center 1 can be explained by the higher number of losses (15 vs. 1 for Center 2 and 9 for Center 3), highlighting a more subtle management issue underlying the screening procedure at Center 1.

That the difference in the number of LF and LD across centers is significative and depends on the particular Level 2 Center is further confirmed by the results of a chi-squared test for independence (based on permutation design, 10,000 permutations) conducted on patients classified as losses (i.e., LD or LF) or no-losses (i.e., IP, TT, NORM) and the particular center (Center 1, 2 or 3). The test result allows us to reject the null hypothesis of independence between the Level 2 centers and their loss/no-loss classification during Level 3 (observed chi-square (χ^2^) = 11.56, DF=2, p-value=.003), indicating a clear relation between the rate of losses and the specific hospital that administered patients during Level 2.

### 2.1 Permanent and temporary hearing loss (Descriptive)

Among the patients who accessed Level 3 in FVG between 2019 and 2023 and received a final diagnosis, 65 were classified as having a permanent hearing impairment (IP), 10 as temporary conductive hearing loss, and 6 as normal hearing subjects. In particular, IP and TT diagnoses were performed during Phase 3D, while normal hearing patients were identified after examinations occurred at Phase 3C (Figure 2).

Available information about these patients and their screening path allows us to investigate the timing of their diagnosis as well as provide details about their clinical conditions. Overall, the majority of patients included in the dataset that accessed Level 3 failed to pass both automated transient evoked otoacoustic emissions (A-TEOAE) and automated auditory brainstem response (A-ABR) examination during Level 1, present JCIH risk factors (JCIH 2019; Beswick et al. 2012), and show a grade of hearing loss (WHO 2021) from mild to moderate in the case of NORM and TT and from mild to profound for IPs, with 47 out of 65 bilaterally.

Regarding the JCIH risk factors specifically (Figure 4), we observe that their presence often led to a refer outcome during Level 1 screening, even though A-TEOAE and A-ABR result negative. The majority (58%) of NORM, TT, and IP patients presented with at least one risk factor (17% multifactorial), while 10% of them had a history of preterm birth. When NORMs and TTs are taken into account, 63% (10 out of 16) of them show at least one risk factor (the most frequent being intensive care of more than 5 days). For IPs, the percentage of patients presenting at least one risk factor decreases to 57% (37 out of 65). Among them, risk factors that occur the most are neonatal intensive care of more than 5 days, family history of hearing loss, presence of syndromes associated with atypical hearing thresholds, and particular birth conditions, such as craniofacial malformations and temporal bone abnormalities.

**Figure 4.**
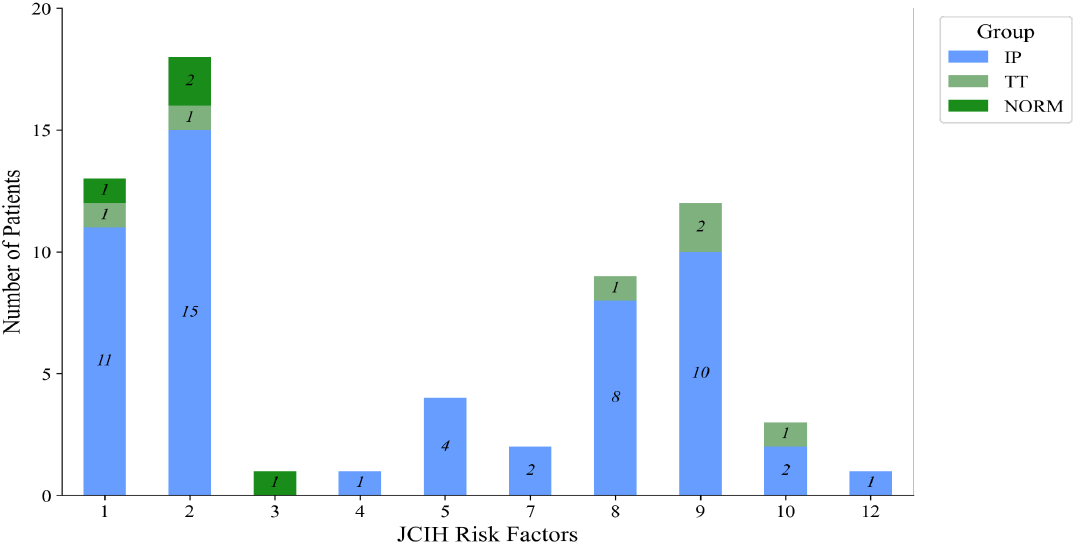
Presence of JCIH risk factors in the different groups, i.e., IP, TT, and NORM. Perinatal risk factors – 1: Family history of early, progressive, or delayed onset permanent childhood hearing loss; 2: Neonatal intensive care of more than 5 days; 3: Hyperbilirubinemia with exchange transfusion regardless of length of stay; 4: Aminoglycoside administration for more than 5 days; 5: Asphyxia or hypoxic ischemic encephalopathy; 6: Extracorporeal membrane oxygenation; 7: In utero infections, such as herpes, rubella, syphilis, and toxoplasmosis, cytomegalovirus; 8: Certain birth conditions or findings (e.g., craniofacial malformations including microtia/atresia, ear dysplasia, oral facial clefting, white forelock, and microphthalmia, congenital microcephaly, congenital or acquired hydrocephalus, temporal bone abnormalities); 9: Syndromes that have been identified with atypical hearing thresholds. Perinatal or postnatal risk factors – 10: Culture-positive infections associated with sensorineural hearing loss, including confirmed bacterial and viral meningitis or encephalitis; 11: Events associated with hearing loss, e.g., significant head trauma, chemotherapy; 12: Caregiver concern regarding hearing, speech, language, developmental delay, and or developmental regression.

With regard to the additional evaluations and examinations carried out during the screening program, the diagnosis of permanent hearing loss was based on a genetic test in 12 patients, and computed tomography or magnetic resonance imaging in 41 cases, to assess the presence of alterations at the level of the inner ear or cerebral abnormalities.. Among those undertaking neuroimaging assessment, 22 show a cerebral alteration, and 2 patients also report an alteration in the inner ear.

Another important aspect to take into account when assessing the overall efficacy of the screening program is the time required to conclude the entire process. As early intervention is essential to address hearing impairments and their consequences, specific recommendations (JCIH 2007, 2019) exist about the optimal timeline of the main stages of the screening, for instance following the 1-3-6 benchmark.

Figures 5a-b report the time interval between birth, the different Phases of Level 3, and prosthesis implantation, both for IPs and TTs. While the protocol of the FVG region adheres to the recommendations of JCIH (2007), e.g., universal hearing screening within 1 month from birth and audiological assessment within 3 months from birth, results indicate a suboptimal timeline on average and difficulties in meeting the 1-2-3 benchmark (JCIH 2019). Indeed, IPs and TTs reach Phase 3A after 6.4 months from birth on average, while Phase 3D occurs after 12 months on average. For what specifically concerns IP’s timeline from birth to implantation, 44% of the patients received audiological assessment and accessed Level 3 of the program within 3 months, as recommended by JCIH guidelines (average 6.3 months), with Phase 3D occurring on average after 11.9 months and implantation after 19 months. All these results are, however, due to a portion of cases achieving Phase 3D and implantation very late (>30 months), while the majority of patients actually receive diagnosis and implantation between 12 to 18 months from birth.

**Figure 5.**
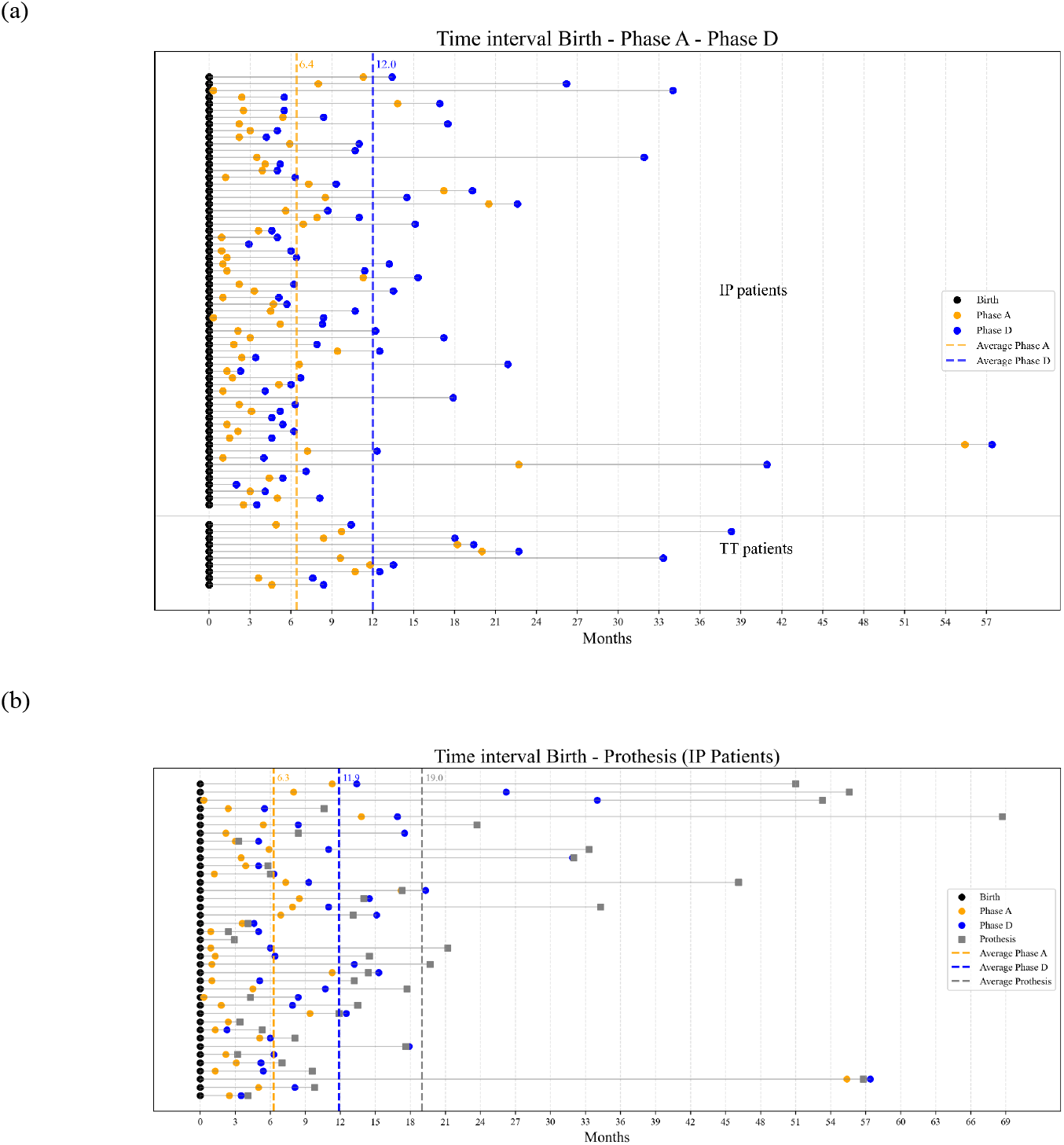
(a) Time interval between birth and Phases 3A and 3D for IP and TT patients. (b) Time interval including prosthesis implantation for IP patients.

Timeline concerns with respect to JCIH guidelines for early intervention are, however, worth examining and might be associated with different factors. Given the focus of the present work, we investigated whether a difference between the timelines of patients diagnosed with temporary and permanent hearing loss exists and, if so, whether this difference is associated with specific organizational and management aspects, such as particular stages of the

### 2.1 Permanent and temporary hearing loss (Statistical analysis)

Different organizational aspects might have an impact on the early assessment of hearing impairments, thus potentially producing cascade effects on further steps of the program, e.g., delayed diagnosis and treatment. For instance, one might hypothesize that issues in the communication and transfer of data between centers involved in the different levels of the program can be associated not only with a higher risk of losses but also with delays and time-related inefficiencies. Another practical aspect worth considering concerns how early in the program a reliable discrimination of the pathological conditions of interest, e.g., temporary or permanent hearing loss, is expected. Indeed, both a high number of patients with suspected hearing losses or a proportion of false positives hard to identify early on might cause inefficiencies in the management, thus leading to errors and delays. These aspects are specifically addressed in the following.

To investigate whether a difference between the timeline associated with permanent and temporary hearing loss is significant and what stages of the screening program have major relevance, we conducted two one-tailed Student’s t-tests on the time intervals occurring between birth and different Level 3 Phases for patients diagnosed with permanent and temporary hearing loss.

In light of the results reported in Fig. 5a, in the first test, we evaluated the hypothesis that the interval occurring between birth and Phase 3A is, on average, greater for TTs than IPs. Given the limited data available, a one-tailed t-test on the time intervals for the two groups was conducted based on a permutation design (10,000 permutations). The test results allows us to reject the null hypothesis that there is no significant difference between the birth to Phase 3A time intervals of TTs and IPs (t(73)=1.68, p-value=.048; 95% CI: [-1.42, 2.53]; Cohen’s d=0.57; 95% CI= [-0.11, 1.25]) and, in particular, to conclude that such a time interval is larger in the case of TTs (TT’s mean interval: 304 days; IP’s mean interval: 174 days). This result demonstrates that time concerns were already involved in the early steps of the screening program, before entering the main Phases of Level 3.

To assess whether Level 3 is also affected in terms of time, a second one-tailed (Welch) t-test was conducted on the time interval between Phase 3A and Phase 3D, corresponding to the final, diagnostic stage of the program. Again, we hypothesized that the time interval was greater for TTs as compared with IPs, and due to the limited number of data, we relied on a permutation design (10,000 rep.).

Test results do not allow us to reject the null hypothesis (t(73)=0.93; p-value=0.12; 95% CI: [-3.53, 1.54]). So, we can conclude that there is no statistically significant difference in the time intervals associated with IPs and TTs when the gap between Phase 3A and Phase 3D is considered.

Therefore, whether a difference in the timeline between temporary and permanent hearing loss exists and is significant, it is in the early steps of the screening procedure, i.e., Levels 1 and 2, with no clear impact of specific stages encountered during Level 3.

The revealed key role of Levels 1 and 2 raises a further question worth addressing, that is, whether these levels of the program involve particular examination strategies that might affect an early and correct discrimination of the two conditions or, for instance, increase the number of false positives, making their identification harder and thus impacting the timeline. In this respect, a central issue regards the particular diagnostic tests employed for early screening during Level 1. At this stage, the adopted protocol requires screening patients based on two objective tests, i.e., A-TEOAE and A-ABR, and then assessing the presence of potential risk factors. Both tests can provide a negative result (pass) or a positive one (refer), and according to the FVG protocol, A-ABR testing is performed subsequently to A-TEOAE, if the result is refer. In particular, the two tests are administered in series, and newborns access Level 2 for audiologic assessment when either (1) both tests result refer or (2) A-TEOAE results refer bilaterally and A-ABR test results pass. Instead, if both test results are pass, but one or more risk factors are present, the patient undergoes an audiologic assessment. Therefore, the particular strategy implemented to aggregate multiple or repeated test results (Cebul et al. 1982; Politser 1982; Fanshawe et al. 2024) and decide whether to refer a patient to Level 2 is particularly relevant for an early and correct discrimination of the conditions of interest, as it might increase the risk of false positive referrals.

Based on the decision three behind Level 1 (Figure 7a), the current strategy to aggregate A-TEOAE and A-ABR tests seems to follow an “all heuristics” or “conjunctive positivity criterion,” based on the Boolean aggregator AND (Böttcher et al. 2025). According to this rule, both tests should result refer, for the outcome of the screening Level 1 to be referred to Level 2 (Figure 7b). The main advantage of such a strategy is to enhance the specificity (or true negative rate) of the aggregated test, when compared to the specificity of the single ones, and to reduce the risk of accepting the hypothesis that the patient is affected by the disease, when in fact the condition is absent (type I error). Therefore, aggregating results of multiple tests via AND operator represents the best strategy to minimize the risk of false positives as compared with other aggregation rules, e.g., OR, or the consideration of a single test result (Böttcher et al. 2025).

**Figure 7.**
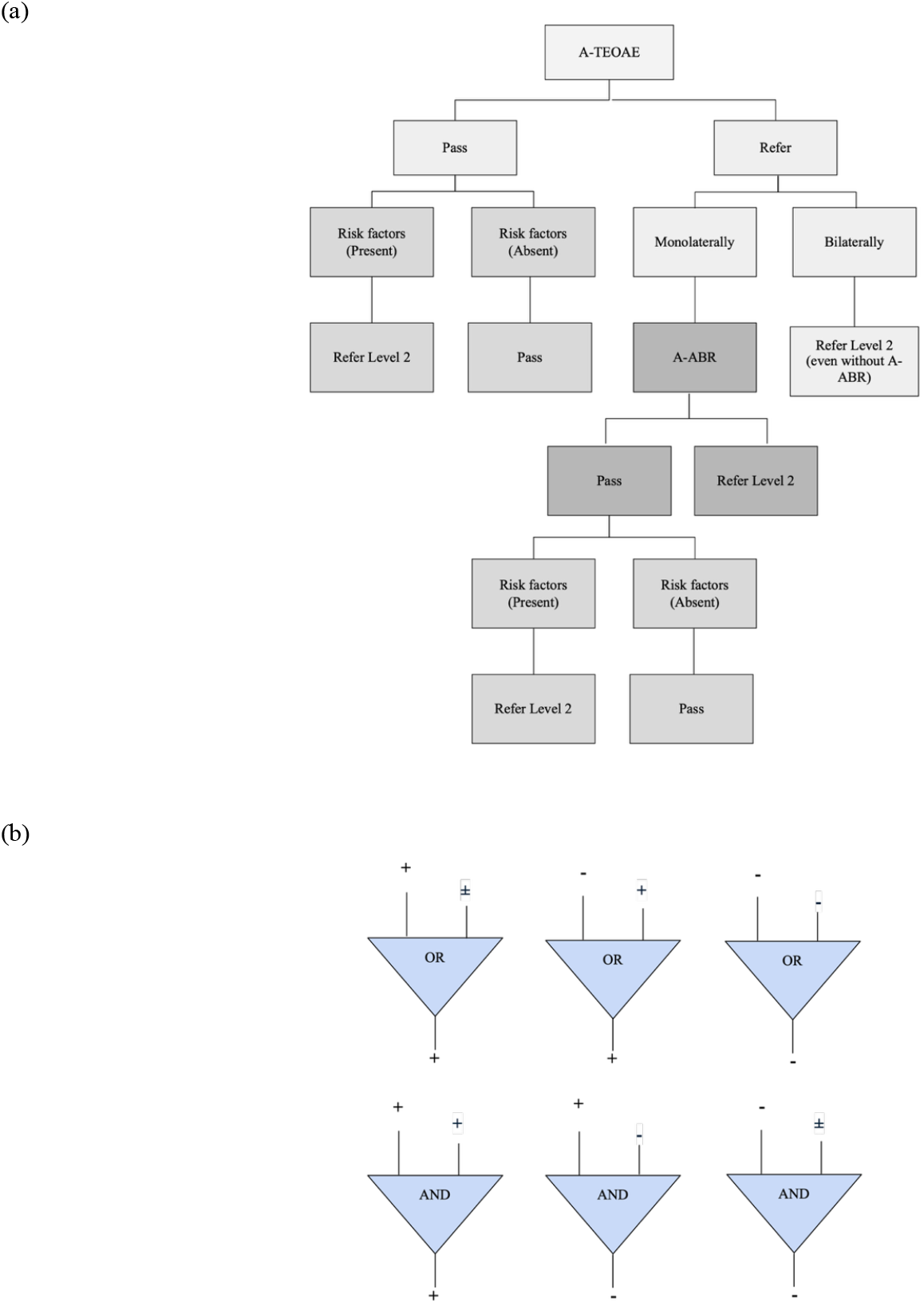
(a) Decision three of Level 1 in 1 FVG based on results of A-TEOAE and A-ABR and the presence of risk factors. (b) Outcomes of AND and OR operators when two input results are aggregated, both in series and parallel testing (in the context of screening, plus and minus signs stand for refer and pass, respectively).

However, the FVG 2019-2023 Database shows that the effective strategy to aggregate results of diagnostic tests performed during Level 1 follows an “any heuristics” or “disjunctive positivity criterion” based on the Boolean aggregator OR. Indeed, the 106 patients arriving at Level 3 generally underwent both tests, even when the A-ABR was not strictly recommended by the FVG protocol, given that a pass result of A-TEOAE or risk factors for which A-ABR is suggested (e.g., intensive care) were absent. Moreover, the dataset includes a few patients who accessed Level 3, even though they passed all Level 1 screening tests and showed no risk factors.

This observation highlights the need to evaluate how results of screening tests are employed during the early step of the program to make decisions (Lin et al. 2007, Benito-Orejas et al. 2008) and, in particular, assessing the effect of using any heuristic rather than the all heuristic as an aggregation strategy for detecting permanent hearing losses. Indeed, in the case of the OR strategy, for a patient to be referred to Level 2, it is sufficient that one of the two administered test results is refer, as illustrated in Figure 7b. This strategy enhances the sensitivity (or true positive rate) of the aggregated examination; that is, the sensitivity of the two tests when considered together is always higher than the sensitivity of each of the two tests taken alone, thus reducing the false negative rate and type II error. However, the main disadvantage of the OR aggregation strategy is that, when compared to single tests, the aggregated examination is less specific, thus increasing the false positive rate and the risk of incurring in type I error (Böttcher et al. 2025).

Based on the available data, we can confirm this fact by computing the actual sensitivities and specificities of A-TEOAE and A-ABR and then the aggregated estimates (based on confusion matrices shown in Tables 3 and 4, see Methods). The effective sensitivity and specificity of A-TEOAE, computed as follows:

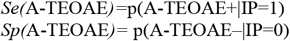

are respectively 0.91 and 0.99. Instead, the sensitivity and specificity of A-ABR

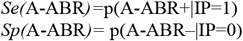

are 0.92 and 0.99. These results confirm a high level of diagnostic accuracy, substantially in line with previous studies that reported sensitivity and specificity above 90-95% for both tests (Sheng et al., 2021; Neumann & Indermark, 2012), with some variation depending on the specific version of the screening instrument used.

**Table 3.** Confusion matrix of A-TEOAE test results.

| A-TEOAE result | IP=1 | IP=0 |
| --- | --- | --- |
| Refer | 59 (TP) | 12 (FP) |
| Pass | 6 (FN) | 39,035 (TN) |

**Table 4.** Confusion matrix of A-ABR test results.

| A-ABR result | IP=1 | IP=0 |
| --- | --- | --- |
| Refer | 59 (TP) | 13 (FP) |
| Pass | 5 (FN) | 39,035 (TN) |

Then, to assess the overall sensitivity (*Se*_*OR*_) and specificity (*Sp*_*OR*_) of Level 1 screening, aggregated estimates can be computed by relying on the OR operator as follows (Böttcher et al. 2025):

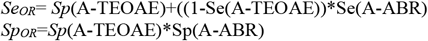

Results show very high estimates, i.e., 0.99 and 0.98, for sensitivity and specificity, respectively. As noted above, the OR aggregation strategy increases sensitivity overall, which was lower for both tests when considered separately. However, the OR aggregation rule always reduces specificity (from 0.99 for both tests to 0.98 for the aggregated case) and, even when very slightly, it in fact increases the risk of having false positives and then the need to manage a higher number of patients during the program with potential consequences on organization and timing.

## Discussion

Between 2019 and 2023, 106 children accessed Level 3 of the neonatal hearing screening program in the FVG Region in Italy. Level 3 is the final stage of the program, involving diagnosis, treatment, and follow-up of hearing losses at the Regional Center for Pediatric Hearing Loss Care (Center 2) in FVG.

The analysis of the children’s reports from identification and diagnosis of the hearing impairments to the prosthesis implantation allows assessing the coverage and efficiency of the procedure as a whole, determining the impact of organizational factors along the different Phases 3A, 3B, 3C, and 3D of the procedure.

While the program is efficiently implemented in the FVG Region since more than ten years and see a coordinated and proficient participations of different actors, such as regional birthing centers, main regional hospitals, and pediatricians, the analyses highlight different management and organizational weaknesses of the current implementation of the program, being related to the number of losses, communication between centers, delays in the treatment of cases, and strategies to aggregate results of early screening test having cascade effects on subsequent phases of the program, such as a greater number of patients (especially, false positives) in need of further examination. The main results of the analysis of the FVG 2019-2023 are subsequently described in order.

First, at the end of the procedure, only 65 of the 106 patients entering Level 3 were effectively diagnosed with a permanent hearing loss, being sensorineural, conductive, or mixed, resulting in an incidence of 1,66‰ in the FVG population. Such a result is due to 10 patients indicated as having a temporary conductive hearing loss, 6 normal hearing conditions, and, most importantly, 28 records have been classified as losses (LDs or LFs). The higher number of losses is mainly observed between Phases 3A and 3B, when patients who have undergone audiological evaluation (Level 2) are referred to Center 2 for diagnosis, treatment, and follow-up. Given that these phases have a clear organizational function and no clinical relevance, the number of losses is a sign of some management weakness concerning these stages of the program. To further investigate the point and rule out the possibility of other explanations, rather than organizational issues, the incidence of patients diagnosed with a permanent hearing loss was compared with the number of total (alive) births managed by each Center. Results reveal that the different birth rates impact the incidence of suspected hearing losses only partially and during the initial steps of the program. A key impact on such a difference is due to management issues, mirrored in the difference in the number of losses associated with each center, which further statistical tests confirm to be significant.

Second, the assessment of the timeline between the different stages of the program highlights that only a percentage of the patients are diagnosed and treated based on the 1-3-6 benchmark recommended by JCIH (2007), while the program does not meet the more recent 1-2-3 guidelines proposed by JCIH (2019). The observed timeline in the FVG region is 1-6-12 on average, considering Phase 3A as a reference for audiological assessment (Level 2) and Phase 3D as indicating the diagnosis and enrollment in intervention services. Such a discrepancy might have different reasons. For instance, the choice of Phase 3A as a reference time point takes into account the transmission of referrals from one center to another. If issues in communicating and transmitting data from Level 2 and Level 3 occur, as observed, the timing of Phase 3A is already affected by potential delays. Moreover, the statistical analysis conducted on the time gap of permanent and temporary hearing loss between birth, Phase 3A, and Phase 3D, reveals that the difference between the time interval in the case of permanent and temporary hearing losses is significant when the birth-Phase 3A interval is considered, but not when the birth-Phase 3D interval is analyzed. These results suggest that different time intervals are associated with different hearing conditions in relation to early stages of the program, i.e., Levels 1 and 2, but not Level 3. Therefore, the suboptimal timing is not associated with delays specifically related to Level 3, but it is largely due to the management of patients during previous steps. Again, probable causes of these delays can be identified in the transmission of referrals and in the early discrimination of the two pathological conditions that, if inefficient, might lead to an increase in the number of patients to be managed because of a high rate of false positives results about permanent hearing impairments. Third, a high rate of false positives is intrinsically related to the quality of the diagnostic tests used during Level 1, in particular. The two important factors to consider in relation to the risks of false positives that reach Level 3 concern the particular diagnostic tests employed and, especially, the strategy to aggregate multiple results during the first part of the screening procedure. During Level 1, newborns are screened with two tests, i.e., A-TEOAE and A-ABR, and assessed for the presence of risk factors. A-ABR measures the auditory evoked potential extracted from ongoing electrical activity in the brain through electrodes placed on the scalp of the baby. Instead, A-TEOAE allows to investigate of the health of the inner ear, based on otoacoustic emissions, that is, sounds generated from the cochlea and transmitted across the middle ear to the external ear canal, where they are recorded. While widely used as an objective diagnostic test for neonatal hearing screening, technical characteristics make A-TEOAE unable to clearly discriminate between conductive and sensorineural hearing loss.

Besides the quality of diagnostic tests, which show a high level of sensitivity and specificity (Sheng et al. 2021; Neumann and Indermark 2012), and their correct implementation, available data reveal that a potential reason for a high number of false positives concerns the specific rule to aggregate test results during Level 1 and decide whether to refer a patient to subsequent evaluation. Indeed, we observe that Level 1 of the screening program implements any heuristic based on OR Boolean operator, according to which, for a patient to be referred to Level 2, it is sufficient that at least one of the administered tests results refer or risk factors are present. While optimal to increase sensitivity with respect to single tests, such a rule decreases the specificity of the aggregated examination, enhancing the risk of incurring in type I error and false positives.

Results reveal that the observed decrease in specificity due to OR aggregation is very slight. However, this strategy necessarily determines an increase in the false positives and of the number of patients to treat. Moreover, when a population of around 39,000 newborns–as in FVG between 2019 and 2023–is taken into account, such a slight increase might have a detrimental effect on the management of suspected hearing losses.

Therefore, even though important for identifying true hearing impairments better than relying on single tests, any heuristics cannot minimize the risk of false positives, potentially causing not only a delayed dismissal of patients from the program, but also time-related inefficiencies in the management of patients with permanent hearing loss. An accurate re-thinking of the best strategies to implement during the early stages of the screening procedure and their potential organizational, economic, and health effects on the program, in particular when multiple test results and clinical outcomes are to be integrated to assess the patients’ conditions and decide how to proceed with treatment, appears to be in need.

## Conclusion

The effectiveness of a hearing screening program cannot be evaluated solely on its clinical performance. Organizational and management aspects play a crucial role in determining the overall quality, scalability, and sustainability of such programs. Screening is inherently a multi-step process that requires careful coordination, timely data transmission, and systematic monitoring. Without robust organizational structures, even clinically sound programs risk inefficiencies, reduced coverage, and delays in care.

The analysis of data from the final stages of the universal newborn hearing screening program implemented in FVG, one of the first Italian regions to adopt such a protocol in 2012, confirms these challenges. The main critical issues concern communication between the different centers involved, such as birthing centers, regional hospitals, pediatricians, and specialized hubs for treatment and follow-up of pediatric hearing loss. These shortcomings may significantly affect program efficiency, leading to both patient losses and delays in intervention.

Importantly, inefficiencies arising in the early stages of the screening pathway can cascade into later phases, making them difficult to address until the very end of the process. For instance, a suboptimal data transmission may influence the timing of early intervention. Moreover, the strategies used to aggregate multiple test results in managing suspected cases immediately after birth may increase the risk of false positives. While a high number of referrals for further audiological evaluations can be justified in the context of screening, it may nevertheless burden the system, reduce efficiency, and raise management and economic concerns. These results underline the importance of carefully balancing diagnostic accuracy with resource management and the need to address these issues directly by health institutes (Orzan et al. 2022). Improving communication between institutions and ensuring standardized monitoring tools are essential not only for optimizing program performance within a single region, but also for strengthening the cooperation between different regions—particularly in cases where patients move between healthcare systems or are referred to specialized institutes outside their area of birth.

In conclusion, the findings of this study suggest that current issues in multi-step neonatal hearing screening programs, such as the one implemented in FVG, to be addressed require careful attention of both the clinical and organizational dimensions. Future solutions should facilitate inter-center communication and support professionals in accessing, sharing, monitoring, and planning patient care in a transparent, standardized, and automated way. In particular, systems and integrated platforms that provide clear oversight of process timing, test protocols, data flow between centers, and aggregation of results for suspected diagnoses are crucial to minimize errors, losses, and delays, ultimately ensuring more effective and equitable programs.

## Methods

### Database

The FVG 2019-2023 Database includes the reports of 106 patients who, between 2019 and 2023, were suspected of having hearing loss and were referred to Level 3 of the neonatal hearing screening procedure in the Friuli-Venezia Giulia Region, Italy. Specifically, the database contains information on the patients’ history across the previous levels of screening, the outcomes of the different audiological examinations they underwent, the diagnosis of hearing impairment, and the subsequent treatment. Moreover, it records the cases of subjects who were dismissed from the program as having normal hearing conditions, as well as those whose evaluations were interrupted or whose documentation was lost (lost to follow-up or lost documentation), thereby providing important details about the overall efficiency of the screening process.

The data were manually extracted from reports available to the personnel of the Institute for Maternal and Child Health IRCCS Burlo Garofolo Trieste, based on entries in a dedicated platform and regular reports from all other regional centers involved in Levels 1 and 2 of the program. Data on the number of births in the FVG region (Table 2) were also available to the Burlo Garofolo Institute, which serves as the FVG regional center administering births.

Overall, the neonatal hearing screening program in FVG is structured across three levels of monitoring for hearing impairments and involves four main stakeholders:

1. Ten birthing centers (BC) in the region, responsible for performing the initial hearing screening test (Level 1) within the first month of life.
2. Family pediatricians, responsible for ongoing hearing surveillance up to the child’s third year of life.
3. Pediatric audiology services located in the region’s three main hospitals and birthing centers (Centers 1, 2, and 3), responsible for conducting comprehensive audiological evaluations (Level 2) for newborns who fail the initial screening or are referred by a pediatrician, and for referring confirmed cases to the Regional Center for Pediatric Hearing Loss Care.
4. The Regional Center for Pediatric Hearing Loss Care, the only one in the FVG region, located at the Burlo Garofolo Institute, responsible for the final audiological and etiological diagnosis, treatment, and follow-up of children with hearing loss (Level 3).

The specific activities and responsibilities of the various operational levels of the Friuli Venezia Giulia program are shown in Figure 1 and described in detail as follows.

During Level 1, newborns are screened at the BC by trained nurses using a two-step procedure. The first test is the automated transient evoked otoacoustic emissions (A-TEOAE), which can be repeated up to two times. If the newborn fails this test, the screening is followed by the automated auditory brainstem response (A-ABR). Both tests are administered to infants considered at risk (see JCIH 2019) within 1 month from birth. The regional health system in FVG has complied with JCIH criteria since the program’s inception. Coverage has consistently exceeded 95%, while the referral rate has remained below 3% since the first year of the program’s formal implementation (Orzan et al. 2021). Family pediatricians are responsible for postnatal hearing surveillance during routine child visits at 1, 3, 6, 9, 12, 18, and 36 months of age. Their tasks include verifying the completion of newborn hearing screening, monitoring age-appropriate auditory and language milestones, and tracking risk indicators for hearing loss.

Children who fail the newborn screening in one or both ears, those lost to or who did not complete the screening process, or those who initially pass but present a congenital or postnatal risk factor as defined by JCIH (2019), are referred to Level 2. At this stage, they undergo a comprehensive audiological evaluation at one of the three pediatric audiology units located in the region’s major hospitals, referred to in this study as Centers 1, 2, and 3. Comprehensive audiological evaluations are performed by expert audiologists—ideally within a single session—and include both objective and subjective tests: otoscopy, diagnostic OAE, tympanometry, ipsilateral acoustic reflexes, and air-conduction ABR to determine hearing thresholds.

Children who fail the Level 2 audiological evaluation are referred to Level 3 for further assessment, diagnosis, and, when necessary, intervention and treatment. The Regional Center for Pediatric Hearing Loss Care, located at the Burlo Garofolo Institute is the only facility in the FVG region responsible for conducting examinations at this final stage of the screening program.

The database analyzed in this study contains detailed information about patients referred to Level 3, allowing us to assess the program’s effective coverage and efficiency along the pathway from identification to diagnosis. Specifically, we examined the number of patients progressing through the subsequent phases of Level 3:

Phase 3A: individuals who failed the second level with a type 3 outcome.

Phase 3B: individuals correctly referred to the third (diagnostic) level.

Phase 3C: individuals who actually presented for evaluation.

Phase 3D: confirmed cases of hearing loss that entered the diagnostic process.

### Statistical analyses

Statistical analyses were conducted in Python using non-parametric permutation tests, due to the limited sample sizes of patient groups. For each analysis, a test statistic (e.g., χ^2^) was computed on the observed data. Observations were then randomly permuted 10,000 times to generate a null distribution of the statistic. P-values were computed by comparing the observed statistic to the null distribution. For significant t tests, the effect size (d’Cohen; Cousineau and Goulet-Pelletier 2021) and the confidence intervals are reported. Confidence intervals were computed using the bootstrap method with N=1000 bootstrap replicas.

All the codes are available at: https://osf.io/5jeby/overview?view_only=d4d53b57f8384489befd32c0c4b525df

### Sensitivity and specificity of diagnostic tests

The effective sensitivity and specificity of diagnostic tests employed during Level 1 of the hearing screening program were computed based on the following confusion matrices. Table 3 compares A-TEOAE results with respect to total births in the FVG region and IP diagnosis. Table 4 reports data for A-ABR results. (A-TEOAE or A-ABR results can be either “Refer,” i.e., positive, or “Pass,” i.e., negative. IP diagnosis can be present, i.e., 1, or absent, i.e., 0. TP= True Positive; FP=False Positive; FN=False Negative; TN=True Negative).

Based on the confusion matrices above, the effective sensitivity (*Se*) and specificity (*Sp*) of the two tests are computed as follows:

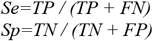

Effective sensitivity (*Se*) and specificity (*Sp*) are then used to compute the aggregated estimates *Se*_*OR*_ and *Sp*_*OR*_ based on the OR operator, as reported in the text.

## Data Availability

Codes and data are available online at https://osf.io/5jeby/overview?view_only=d4d53b57f8384489befd32c0c4b525df.

https://osf.io/5jeby/overview?view_only=d4d53b57f8384489befd32c0c4b525df

